# Sex Differences in the Incidence and Risk of Ankle-Foot Complex Stress Fractures Among U.S. Military Personnel

**DOI:** 10.1101/2021.05.14.21254379

**Authors:** Andrew J. MacGregor, Sarah A. Fogleman, Amber L. Dougherty, Camille P. Ryans, Cory F. Janney, John J. Fraser

## Abstract

**Background:** The objective of this study was to evaluate sex differences in the incidence and risk of ankle-foot complex (AFC) stress fractures among U.S. military personnel, which could assist in developing management strategies as females assume a greater role in U.S. military operations.

**Methods:** The Defense Medical Epidemiological Database was used to identify all diagnosed AFC stress fractures in military personnel from 2006 to 2015. Cumulative incidence of AFC stress fractures was calculated and compared by year, service branch, and military rank. Sex differences in the risk of AFC stress fractures by occupation were examined, and integrated (i.e., male and female) occupations were compared with non-integrated (i.e., male-only) occupations.

**Results:** A total of 43,990 AFC stress fractures were identified. The overall incidence rate was 2.76 per 1,000 person-years for males and 5.78 per 1,000 person-years for females. Females consistently had higher incidence of AFC stress fractures across all subgroups, particularly among enlisted personnel. Female enlisted service members had the highest risk of AFC stress fractures in aviation [relative risk (RR) = 5.74; 95% confidence interval [CI] 4.80–6.87] and artillery/gunnery (RR = 5.15; 95% CI 4.62–5.75) occupations. Females in integrated occupations had significantly higher rates of AFC stress fractures than males in both integrated and non-integrated occupations (i.e., special forces, infantry, and mechanized/armor).

**Conclusions:** Females in the U.S. military have a higher risk of AFC stress fractures than males. As integration of females into previously sex-restricted occupations continues, focused prevention efforts may be needed to reduce injury burden and maximize medical readiness.

## Introduction

Lower extremity stress fractures occur when repetitive microtrauma, typically from overuse, causes a partial or complete fracture of the bone.^1^ These injuries are most prevalent in populations with vigorous physical activity, such as athletes and military personnel.^2^ Military recruits in basic training represent a vulnerable population for musculoskeletal injuries, and one epidemiological study found that 4% of Marine Corps recruits sustained a stress fracture during boot camp.^1–3^ This is significantly higher than the 0.5% prevalence in the general population.^2,4^ Service members in non-training environments also experience lower extremity stress fractures, likely due to the physical nature of their occupational duties and exercise regimens required to maintain readiness.^1^ Increased body mass, which is becoming more problematic in both recruits and service members,^5,6^ is a strong risk factor for stress fracture.^7^ This is particularly concerning given that geopolitical shifts may necessitate the recruitment of individuals that do not meet current body composition standards, especially during periods of force growth for conflict, which could lead to an increased risk and burden of stress fracture in the military.^8^

Stress fractures are a significant problem in the military and present numerous challenges. Waterman and colleagues^9^ used diagnostic codes to identify lower extremity stress fractures in the military between 2009 and 2012, and found an overall incidence rate of 5.69 per 1,000 person-years. In this study, more than half of the stress fractures occurred in the ankle-foot complex (AFC), which consists of the tibia, fibula, and metatarsals, and a majority of all stress fractures were among junior enlisted personnel in the Army and Marine Corps. Importantly, because younger military personnel account for most of the forces deployed to combat, these stress fractures can threaten operational readiness.^10^ Additional concerns of lower extremity stress fractures in the military include lost duty time and increased discharge rates, which present serious financial and manpower implications.^2,11,12^ Moreover, a recent military study found that injuries to the ankle and foot were one of the primary reasons for limited duty profiles (i.e., inability to fully engage in required physical tasks), which further highlights the impact of AFC stress fractures.^13^

Sex is a nonmodifiable risk factor for AFC and other lower extremity stress fractures, and military studies show female service members have greater risk than males.^1,2,14,15^ Two studies that assessed military training populations found that females had more than 3-times higher risk of stress fractures than their male counterparts.^14,15^ A similar association was found among the overall active duty population.^9^ Posited mechanisms for the higher risk in females include disparities in physical fitness levels, anatomical differences (e.g., skeletal structure and density) that affect biomechanics, and endocrinological factors.^1,2,9^ Importantly, none of the aforementioned studies jointly examined sex and military occupation. With the diverse range of specialized military occupations that require differing levels of physical rigor,^16^ and operational environments that may expose service members to hazards associated with AFC stress fracture, there is a need to investigate sex-related differences by occupation.

Women have seen increased utilization in U.S. military operations over the years, making the study of sex differences altogether more important.^17,18^ The assignment of women to specific military occupations has gained national attention. Recently, the U.S. Secretary of Defense opened all occupations to women who met validated occupational standards, including frontline combat units that were previously restricted to men.^19^ Further, research on women’s health issues has become a priority in the military.^20^ It is therefore essential to evaluate sex differences in AFC stress fractures by military occupation to establish baselines for preventive strategies.

The objective of this study was to evaluate sex differences in overall incidence of AFC stress fractures among U.S. military personnel between 2006 and 2015, and to assess risk by occupation. More specifically, this study aimed to: (1) describe the rates of AFC stress fractures among females and males in the military by year, rank, and service branch; (2) evaluate sex differences in AFC stress fracture risk among specific military occupations; and (3) compare AFC stress fracture rates for integrated occupations with those that were not integrated during the study period.

## Patients and Methods

A population-based epidemiological retrospective study of all service members in the U.S. Armed Forces was performed to assess sex differences in AFC stress fracture incidence between 2006 and 2015. The Defense Medical Epidemiological Database (DMED) was utilized to identify health care encounters.^21^ This database provides aggregated data for *International Classification of Diseases, Ninth Revision, Clinical Modification* (ICD-9-CM) codes and de-identified patient characteristics, including sex, categories of military occupations, and branch of service for all active duty and reserve military service members.^22^ The DMED is HIPAA compliant and has been used previously in epidemiological studies of lower extremity injury in the military.^9,23,24^ This study was approved as non–human-subjects research by the Institutional Review Board at Naval Health Research Center, San Diego, CA.

The database was queried for individuals with AFC stress fractures, defined as a primary diagnosis of stress fracture of the tibia or fibula (ICD-9-CM code 733.93) or metatarsals (733.94) on the initial medical encounter. These are the only diagnostic codes for stress fractures specific to the AFC. Those with repeat visits for the same diagnosis were only counted once in all analyses. Variables indicating sex, service branch (Army, Navy, Marine Corps, or Air Force), rank (enlisted or officer), and military occupation were obtained from DMED. Population at risk was derived by DMED for each year to obtain an estimate of person-years (p-y).

Occupation was classified separately for enlisted personnel and officers. Enlisted occupations included: special forces; infantry; mechanized/armor; artillery/gunnery (e.g., general artillery, missile artillery, rocket artillery); aviation (e.g., air crew, pilots, navigators); engineers; maintenance (e.g., craftworkers, electric/electronic/mechanical repair); administration/intelligence/communication; logistics (e.g., service and supply handlers); maritime/naval specialties (e.g., seamen, boat operators); and training (e.g., cadets and officer candidates). Officer occupations included: ground and naval gunfire; aviation; engineering and maintenance; administration; operations and intelligence; logistics; services (e.g., health care and science); and training. All occupations contained both male and female service members except for the following male-only enlisted occupations: special forces, infantry, and mechanized/armor. There were no male-only officer occupations. These male-only enlisted occupations were termed “non-integrated” for the purposes of this study, whereas all other enlisted occupations containing both sexes were termed “integrated.”

### Statistical analyses

Cumulative incidence of patients diagnosed with AFC stress fractures were calculated and compared by year for male and female service members, enlisted and officers, and each service branch. Chi-square or Fisher’s exact statistics and corresponding p-values were used for comparison purposes. Relative risk (RR) point estimates and 95% confidence intervals (CIs) were calculated in the assessment of sex and occupation for integrated occupations. Attributable risk (AR) was calculated as an indicator of the impact of sex on the analysis of occupation (i.e., percent of risk attributable to female sex). Incidence of AFC stress fractures in non-integrated enlisted occupations were compared with integrated enlisted occupations using Chi-square statistics and p-values. An alpha level of 0.05 was used. All analyses were performed using Microsoft Excel for Mac 2016 (Microsoft Corp., Redmond, WA) and OpenEpi.^25^

## Results

A total of 43,990 AFC stress fractures were identified in military personnel during the study period. The overall incidence of AFC stress fractures during the study period was 3.20 per 1,000 p-y, with 2.76 per 1,000 p-y for males (32,388 fractures and 11,731,979 p-y) and 5.78 per 1,000 p-y for females (11,602 fractures and 2,007,049 p-y). Table 1 details the sex-specific cumulative incidence of AFC stress fractures stratified by year, rank, and service branch. Across all years, the highest rates of AFC stress fractures were among enlisted, females, and Army and Marine Corps personnel. Specifically, the largest incidence of AFC stress fractures was among enlisted females in the Marine Corps (11.8 per 1,000 p-y compared with 4.8 per 1,000 p-y for males) and Army (8.6 per 1,000 p-y compared with 4.0 per 1,000 p-y for males). The largest relative difference between females and males was among enlisted Navy personnel, with females experiencing more than 3-times higher incidence of AFC stress fractures than males (5.8 per 1,000 p-y vs. 1.6 per 1,000 p-y). Within all individual years and service branches, enlisted females had a significantly higher incidence of AFC stress fracture than enlisted males (all *p*s < 0.001). The largest cumulative incidence for enlisted males and females was in the Marine Corps in 2006 (23.2 per 1,000 p-y for females and 7.9 per 1,000 p-y for males) and 2007 (12.8 per 1,000 p-y for females and 7.4 per 1,000 p-y for males). Among officers, incidence of AFC stress fractures was mostly higher for females, though statistical significance was inconsistent within individual years. The largest sex difference among officers was seen in the Marine Corps, with an incidence of 5.5 per 1,000 p-y for females and 2.0 per 1,000 p-y for males. Similar to female enlisted personnel, female officers had the highest incidence in the Marine Corps during the years 2006 (16.9 per 1,000 p-y) and 2007 (11.6 per 1,000 p-y).

**TABLE 1.**
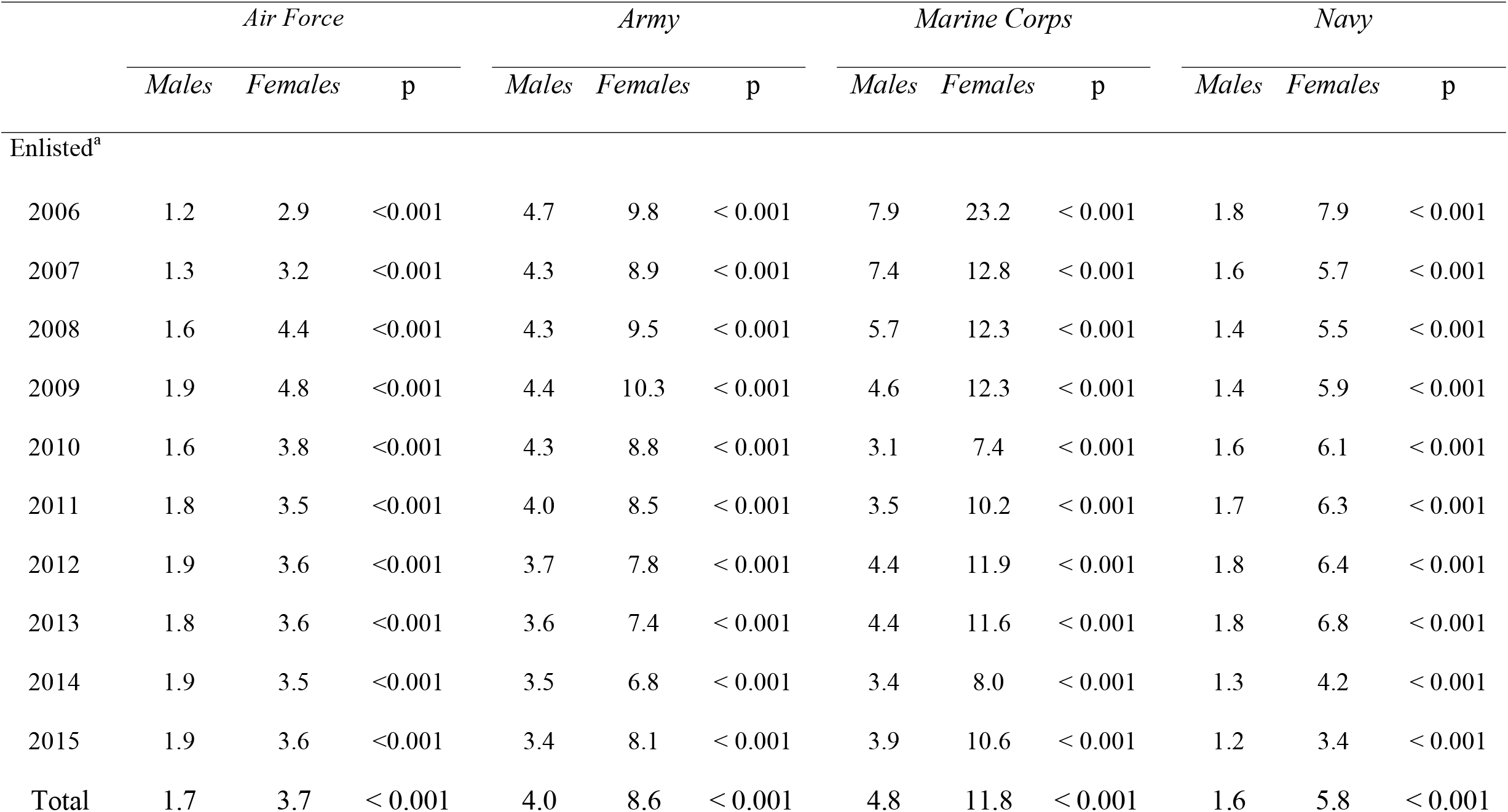
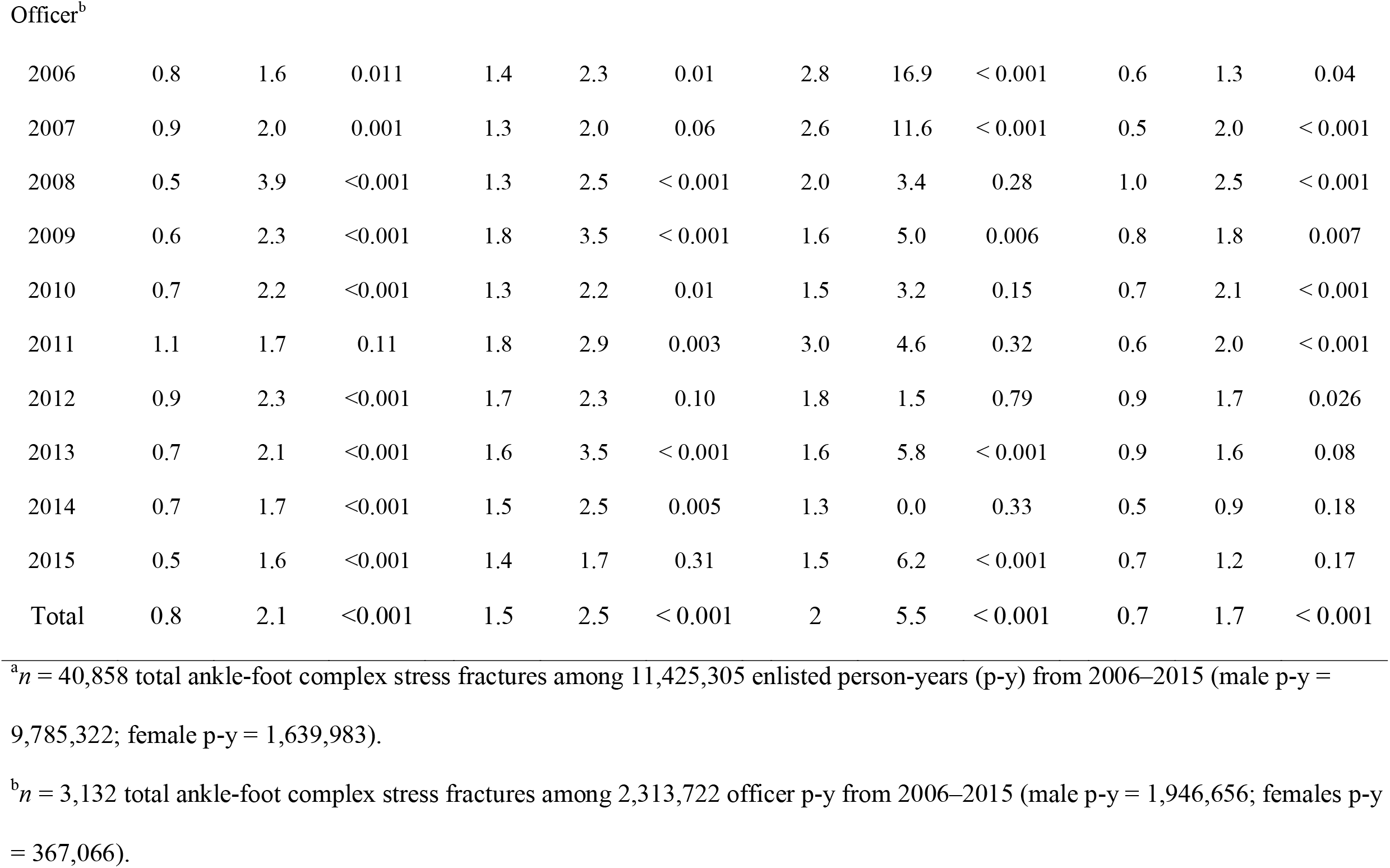
Sex-Specific Incidence Rates (Per 1,000 Person-Years) of Ankle-Foot Complex Stress Fractures, U.S. Military Personnel, 2006–2015.

|  | *Air Force* |  |  | *Army* |  |  | *Marine Corps* |  |  | *Navy* |  |  |
| --- | --- | --- | --- | --- | --- | --- | --- | --- | --- | --- | --- | --- |
|  | *Males* | *Females* | *p* | *Males* | *Females* | *p* | *Males* | *Females* | *p* | *Males* | *Females* | *p* |
| Enlisted<sup>a</sup> |  |  |  |  |  |  |  |  |  |  |  |  |
| 2006 | 1.2 | 2.9 | <0.001 | 4.7 | 9.8 | < 0.001 | 7.9 | 23.2 | < 0.001 | 1.8 | 7.9 | < 0.001 |
| 2007 | 1.3 | 3.2 | <0.001 | 4.3 | 8.9 | < 0.001 | 7.4 | 12.8 | < 0.001 | 1.6 | 5.7 | < 0.001 |
| 2008 | 1.6 | 4.4 | <0.001 | 4.3 | 9.5 | < 0.001 | 5.7 | 12.3 | < 0.001 | 1.4 | 5.5 | < 0.001 |
| 2009 | 1.9 | 4.8 | <0.001 | 4.4 | 10.3 | < 0.001 | 4.6 | 12.3 | < 0.001 | 1.4 | 5.9 | < 0.001 |
| 2010 | 1.6 | 3.8 | <0.001 | 4.3 | 8.8 | < 0.001 | 3.1 | 7.4 | < 0.001 | 1.6 | 6.1 | < 0.001 |
| 2011 | 1.8 | 3.5 | <0.001 | 4.0 | 8.5 | < 0.001 | 3.5 | 10.2 | < 0.001 | 1.7 | 6.3 | < 0.001 |
| 2012 | 1.9 | 3.6 | <0.001 | 3.7 | 7.8 | < 0.001 | 4.4 | 11.9 | < 0.001 | 1.8 | 6.4 | < 0.001 |
| 2013 | 1.8 | 3.6 | <0.001 | 3.6 | 7.4 | < 0.001 | 4.4 | 11.6 | < 0.001 | 1.8 | 6.8 | < 0.001 |
| 2014 | 1.9 | 3.5 | <0.001 | 3.5 | 6.8 | < 0.001 | 3.4 | 8.0 | < 0.001 | 1.3 | 4.2 | < 0.001 |
| 2015 | 1.9 | 3.6 | <0.001 | 3.4 | 8.1 | < 0.001 | 3.9 | 10.6 | < 0.001 | 1.2 | 3.4 | < 0.001 |
| Total | 1.7 | 3.7 | < 0.001 | 4.0 | 8.6 | < 0.001 | 4.8 | 11.8 | < 0.001 | 1.6 | 5.8 | < 0.001 |
| Officer<sup>b</sup> |  |  |  |  |  |  |  |  |  |  |  |  |
| 2006 | 0.8 | 1.6 | 0.011 | 1.4 | 2.3 | 0.01 | 2.8 | 16.9 | < 0.001 | 0.6 | 1.3 | 0.04 |
| 2007 | 0.9 | 2.0 | 0.001 | 1.3 | 2.0 | 0.06 | 2.6 | 11.6 | < 0.001 | 0.5 | 2.0 | < 0.001 |
| 2008 | 0.5 | 3.9 | <0.001 | 1.3 | 2.5 | < 0.001 | 2.0 | 3.4 | 0.28 | 1.0 | 2.5 | < 0.001 |
| 2009 | 0.6 | 2.3 | <0.001 | 1.8 | 3.5 | < 0.001 | 1.6 | 5.0 | 0.006 | 0.8 | 1.8 | 0.007 |
| 2010 | 0.7 | 2.2 | <0.001 | 1.3 | 2.2 | 0.01 | 1.5 | 3.2 | 0.15 | 0.7 | 2.1 | < 0.001 |
| 2011 | 1.1 | 1.7 | 0.11 | 1.8 | 2.9 | 0.003 | 3.0 | 4.6 | 0.32 | 0.6 | 2.0 | < 0.001 |
| 2012 | 0.9 | 2.3 | <0.001 | 1.7 | 2.3 | 0.10 | 1.8 | 1.5 | 0.79 | 0.9 | 1.7 | 0.026 |
| 2013 | 0.7 | 2.1 | <0.001 | 1.6 | 3.5 | < 0.001 | 1.6 | 5.8 | < 0.001 | 0.9 | 1.6 | 0.08 |
| 2014 | 0.7 | 1.7 | <0.001 | 1.5 | 2.5 | 0.005 | 1.3 | 0.0 | 0.33 | 0.5 | 0.9 | 0.18 |
| 2015 | 0.5 | 1.6 | <0.001 | 1.4 | 1.7 | 0.31 | 1.5 | 6.2 | < 0.001 | 0.7 | 1.2 | 0.17 |
| Total | 0.8 | 2.1 | <0.001 | 1.5 | 2.5 | < 0.001 | 2 | 5.5 | < 0.001 | 0.7 | 1.7 | < 0.001 |

<sup>a</sup> $n = 40,858$ total ankle-foot complex stress fractures among 11,425,305 enlisted person-years (p-y) from 2006–2015 (male p-y = 9,785,322; female p-y = 1,639,983).
<sup>b</sup> $n = 3,132$ total ankle-foot complex stress fractures among 2,313,722 officer p-y from 2006–2015 (male p-y = 1,946,656; females p-y = 367,066).

Sex differences in risk of AFC stress fractures by integrated military occupations are shown in Table 2. In 15 of the 16 occupations, females relative to males had a significantly higher risk of AFC stress fractures. The largest magnitude of risk was seen in the aviation (RR = 5.74; 95% CI 4.80–6.87) and artillery/gunnery (RR = 5.15; 95% CI 4.62–5.75) enlisted occupations, which also had the largest AR percentages (82.6% and 80.6%, respectively). The only occupation that was not statistically different by sex was among officers in the ground and naval gunfire occupation (RR = 0.95; 95% CI 0.61–1.49), though this occupation also had the lowest number of AFC stress fractures among females during the study period (*n* = 20). All other integrated military occupations had a significantly higher risk of AFC stress fractures among females than males with RRs ranging from 1.79-3.74.

**TABLE 2.** Relative Risk and Attributable Risk of Ankle-Foot Complex Stress Fractures for Females Versus Males in Integrated Occupations, U.S. Military Personnel, 2006–2015.

| *Occupation* | *RR (95% CI)* | *AR%* | *p* |
| --- | --- | --- | --- |
| Enlisted |  |  |  |
| Artillery/gunnery | 5.15 (4.62–5.75) | 80.6 | < 0.001 |
| Aviation | 5.74 (4.80–6.87) | 82.6 | < 0.001 |
| Engineers | 3.16 (2.56–3.90) | 68.3 | < 0.001 |
| Maintenance | 2.91 (2.75–3.08) | 65.6 | < 0.001 |
| Administration/intelligence/communication | 1.92 (1.83–2.02) | 48.0 | < 0.001 |
| Logistics | 2.87 (2.67–3.08) | 65.1 | < 0.001 |
| Maritime/naval specialties | 2.41 (2.04–3.08) | 58.5 | < 0.001 |
| Training | 3.74 (2.87–4.87) | 73.3 | < 0.001 |
| Officer |  |  |  |
| Ground and naval gunfire | 0.95 (0.61–1.49) | -4.8 | 0.84 |
| Aviation | 2.33 (1.52–3.55) | 57.0 | < 0.001 |
| Engineering and maintenance | 2.14 (1.71–2.68) | 53.3 | < 0.001 |
| Administration | 1.95 (1.51–2.51) | 48.6 | < 0.001 |
| Operations and intelligence | 2.27 (1.75–2.96) | 56.0 | < 0.001 |
| Logistics | 2.28 (1.77–2.94) | 56.1 | < 0.001 |
| Services | 2.08 (1.80–2.40) | 52.0 | < 0.001 |
| Training | 1.79 (1.33–2.40) | 44.1 | < 0.001 |

AR, attributable risk; CI, confidence interval; RR, relative risk.

Table 3 presents a comparison of incidence rates of AFC stress fractures for integrated versus non-integrated enlisted military occupations. The most populous non-integrated occupation was infantry, contributing 84.1% (966,991 of 1,150,444) of the total p-y. Males in special operations (0.8 per 1,000 p-y) and mechanized/armor (2.4 per 1,000 p-y) had significantly lower rates of AFC stress fracture than those in integrated occupations (2.9 per 1,000 p-y), whereas infantry (5.0 per 1,000 p-y) had significantly higher rates (*p*s ≤ 0.001). All three of the non-integrated occupations had significantly lower rates of AFC stress fracture compared with females (6.6 per 1,000 p-y) in integrated occupations (*p*s < 0.001).

**TABLE 3.**
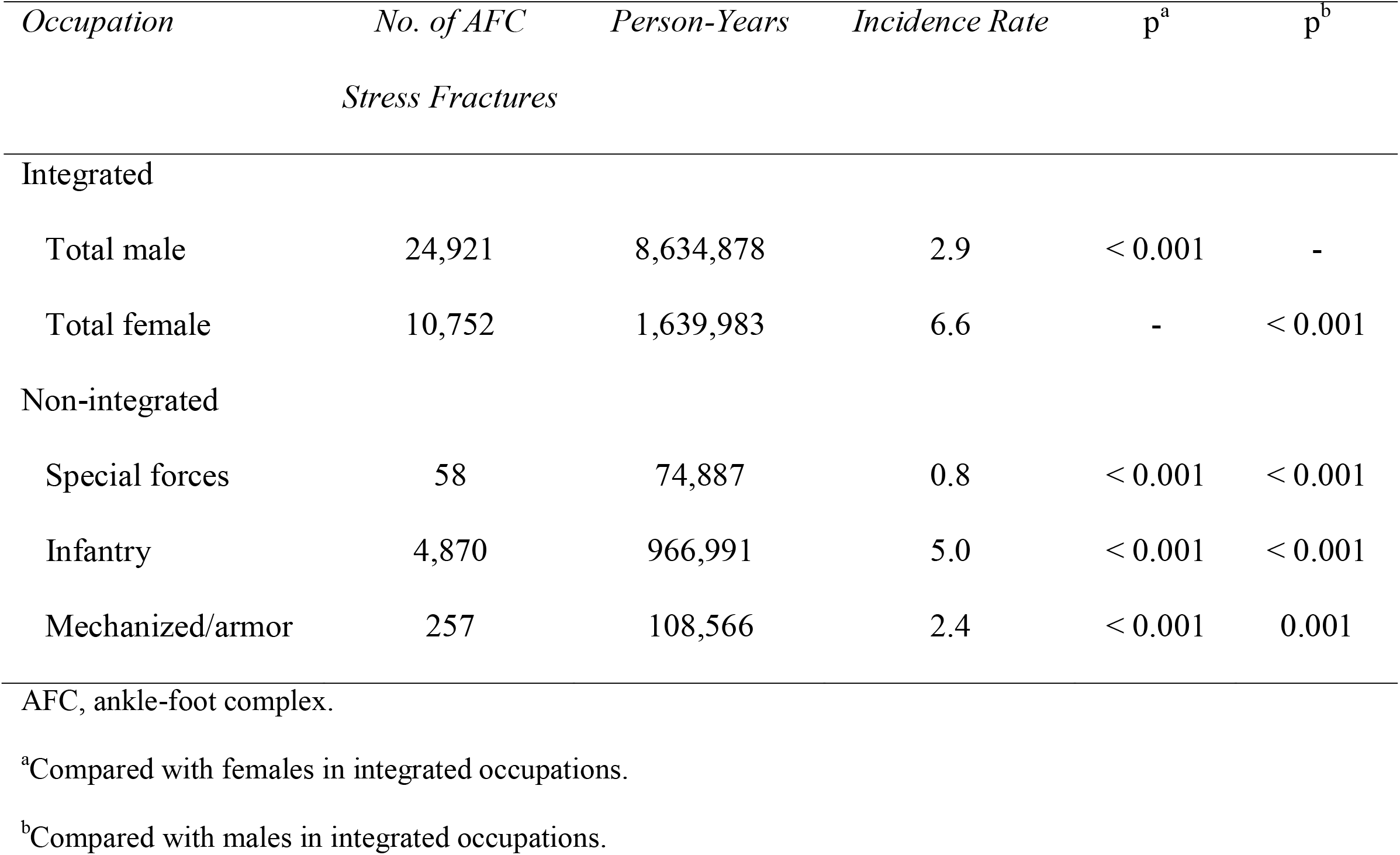
Incidence Rates (Per 1,000 Person-Years) of Ankle-Foot Complex Stress Fractures in Integrated Versus Non-Integrated Enlisted Occupations, U.S. Military Personnel, 2006–2015.

## Discussion

Stress fractures among military personnel are known to occur at higher rates in females compared with males.^1,2,14,15^ To our knowledge, this study is the first to specifically address AFC stress fractures and to examine sex differences by year, service, and occupation. We found that sex differences in the annual incidence of AFC stress fractures were mostly consistent among enlisted personnel from 2006 to 2015, but less consistent among officers. Similar to previous research on lower extremity stress fractures,^9^ females in the Army and Marine Corps had the greatest burden of AFC stress fractures. Certain occupations showed a more profound sex difference in AFC stress fracture risk, and the rate among females in integrated occupations was higher than that of males in both integrated and non-integrated occupations. As women assume an expanded role in future combat operations, they may be at greater risk of AFC stress fractures, and military medical leadership should be appropriately prepared.

Across enlisted occupations, females had consistently higher rates of AFC stress fractures than males for all years of the study period and all branches of service. Sex differences were less apparent among officers, likely due to a smaller sample size and training or occupational duties that were less physically demanding than for enlisted personnel. Because officers have greater autonomy in deciding the mode and frequency of physical training when compared with their enlisted counterparts, it is highly plausible that they also have greater control in adjusting their training if symptoms manifested during the pre-clinical phase of injury. Self-management early in the injury course through cross-training, timing, and volume of training may prevent injury progression and eliminate the necessity to seek medical care. This may also be true for more senior enlisted members, as Waterman et al.^9^ found that the risk of stress fracture was lower among service members in senior compared with junior enlisted ranks. A closer examination of rank may be warranted in subsequent studies.

The highest rates of AFC stress fracture occurred among female Marines in 2006, which could be explained by the operational tempo at that time. Approximately 1 in 4 Marines experienced multiple combat deployments in this period when U.S. combat casualties were peaking during the conflict in Iraq and troop levels surged.^10,26,27^ Operational tempo may also explain why AFC stress fracture rates were highest in male and female members of the Army and Marines, as these two services accounted for most of the ground combat deployments during the Iraq conflict.^28^ More research is needed to determine how changes in operational tempo during active military conflict modifies the risk of musculoskeletal injuries with respect to deployments and pre-deployment training cycles.

Altogether, Marines had the highest rate of AFC stress fractures and this could reflect differences in fitness standards across services, as Marines have the most rigorous physical readiness testing.^29^ Enlisted Navy personnel yielded the largest magnitude sex difference, with the rates of AFC stress fractures more than 3-times higher in females than males. This may be due to unique hazards in the shipboard environment, such as ship movement and non-pliable steel decks that affect kinetics during function, which likely contributes to musculoskeletal injury rates among Navy personnel, though it is not known whether these hazards are potentiated by sex differences.^30,31^ It may also be attributed to higher body mass in Navy personnel compared with other services.^32,33^ Longitudinal studies are needed to further elucidate the etiology of sex differences in AFC stress fractures and possible areas where interventions could be targeted (e.g., equipment, ergonomics).

Overall, the artillery/gunnery and aviation enlisted occupations had the greatest sex differences in rates in AFC stress fracture, which were more than 5-times higher for females than males. Specific duties and tasks in the artillery/gunnery occupations, such as heavy load carriage, may be a factor. Previous studies have found that load carriage is a correlate of musculoskeletal conditions, particularly of the foot,^34,35^ and a study among Australian military members found that female soldiers had more load carriage injuries to the foot than male soldiers.^36^ Although no military studies have examined injuries among aviation workers, civilian research points to musculoskeletal issues in this occupational subgroup that may be mediated by stress.^37,38^ It is difficult to determine the cause of occupational disparities in AFC stress fractures without specific information on the tasks performed by each occupation. Future research should incorporate task analysis when comparing the risk of these injuries by sex and occupation to facilitate the development of interventions that could mitigate injury risk.

The findings comparing integrated with non-integrated occupations provides insight regarding the long-term burden of AFC stress fractures and presents an opportunity to proactively plan and implement interventions and medical services. The relative risks for females compared with males in all integrated enlisted occupations ranged from 1.92 to 5.74, which could translate to nearly double the risk or higher for females when fully integrated in infantry, special operations, and mechanized/armor occupations. This aligns with a recent study examining the impact of integrated combat units, where the rate of musculoskeletal injuries was more than 2-times higher in females than males.^39^ The greatest impact would be seen among infantry, which is the largest of these previously non-integrated occupations that also had the highest rate of AFC stress fracture. Injury prevention programs may need to target to these occupations, particularly during times of high operational tempo. To prevent adverse impacts on operations, surveillance programs should continue to monitor injury incidence among military populations with a focus on sex differences in newly integrated occupations. The findings of this study can also be used to refine medical capabilities, to include clinicians with the knowledge, skills, and abilities to effectively provide evaluation and treatment following injury onset.^8^

This study has several strengths. The utilization of the DMED allowed for examination of population-level sex differences in AFC stress fractures across several strata, including military rank, branch of service, year, and occupation. Most notably, the large range of military occupations in the present study has not been previously examined in stress fracture research. Further, the use of person-years allowed for a more accurate assessment of population at risk. There are also limitations that warrant mention. In contrast to Waterman et al.,^9^ the unspecified ICD-9-CM code 733.95 indicating “stress fracture of other bone” was excluded for the purpose of specificity, which may have underestimated the AFC stress fracture burden if some fractures (e.g., calcaneus, talus) were classified using the nonspecific code. Further, using diagnostic codes has inherent weaknesses, especially in military populations where there are many determinants that can influence care-seeking,^8^ and the data used in this study only represent individuals who sought treatment for their injuries. In addition, we were unable to account for individuals with recurrent AFC stress fractures. Finally, although personnel were categorized by their military occupation, a detailed task analysis to determine the specific duties that may be associated with these injuries was not possible.

## Conclusion

The role of women in the military is increasing, and by the time of the next U.S. conflict, several occupations that previously restricted women may be fully integrated. The findings of the present study assist in establishing the burden of AFC stress fractures, which is essential for the development of targeted preventive health programs. We found a consistently higher rate of AFC stress fractures among female service members compared with males that was more apparent among enlisted personnel. Future studies should define the key etiological factors of AFC stress fractures in the military, including a detailed task analysis to determine why sex differences are more profound in certain occupations, and identify areas of intervention as women integrate into combat occupations. The continued study of sex differences is paramount to address the changing demographics of the military.

## Data Availability

The data that support the findings of this study are available from the corresponding author upon reasonable request.

## Authors Contributions

A.J.M. and J.J.F. designed the study and drafted the article. J.J.F. acquired the data. A.J.M. and J.J.F. analyzed the data. All authors interpreted the data, critically revised the manuscript for important intellectual content, and approved the final version to be published.

## Author Disclosure Statement

No competing financial interests exist.

## Funding Statement

No external funding was received for this work.

---

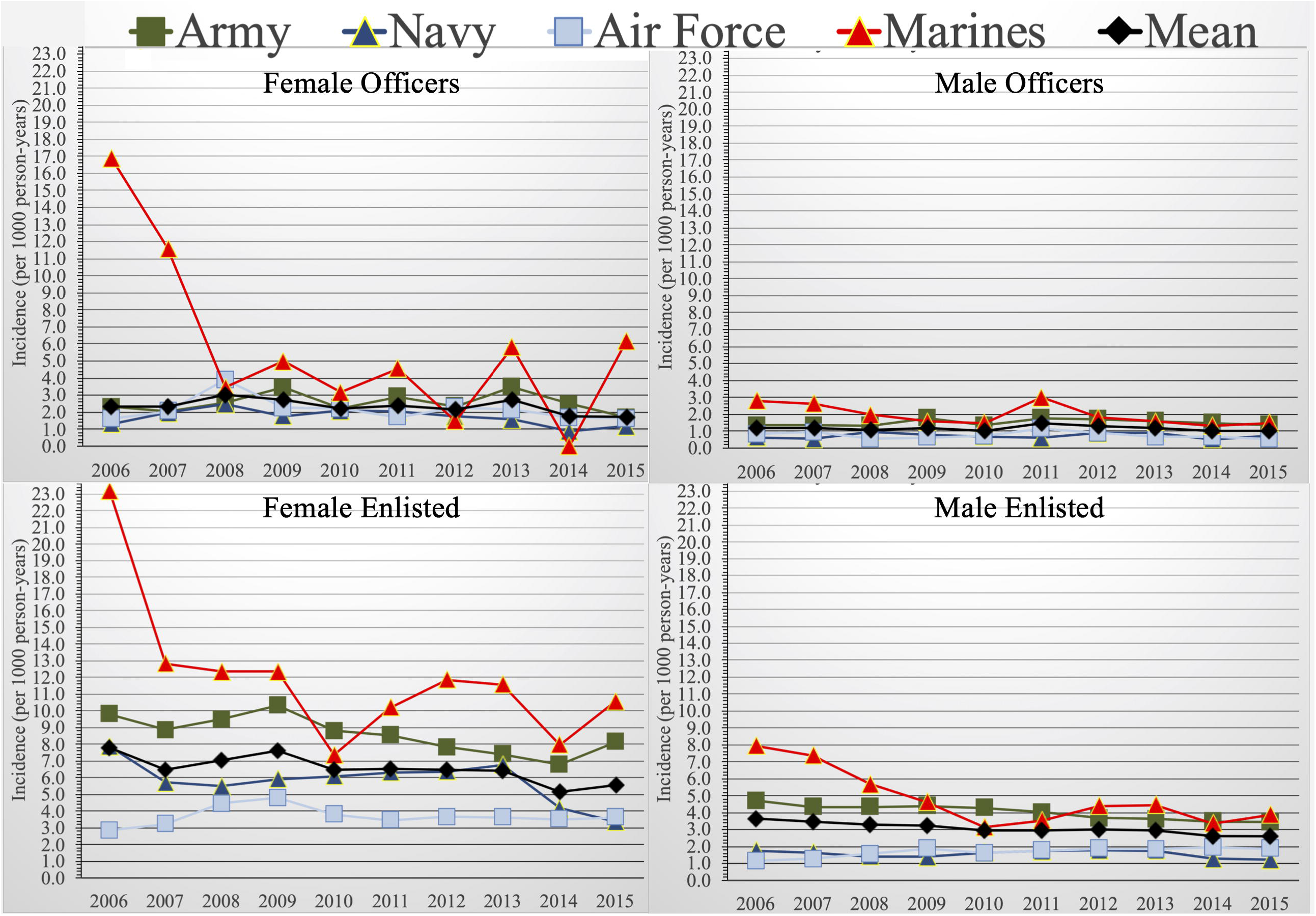

## Notes

### Competing Interest Statement

The authors have declared no competing interest.

### Funding Statement

The authors are military service members or employees of the U.S. Government. This work was prepared as part of their official duties.

### Author Declarations

The study protocol was approved by the Naval Health Research Center Institutional Review Board in compliance with all applicable Federal regulations governing the protection of human subjects. Research data were derived from an approved Naval Health Research Center Institutional Review Board protocol, number NHRC.2020.0205-NHSR.

## References

1. Jacobs JM, Cameron KL, Bojescul JA. Lower extremity stress fractures in the military. Clin Sports Med. 2014 Oct;33(4):591–613.

2. Gun BK, McCoy AC, Wang KC, Waterman BR. Stress fractures: Lessons from military research. Available at https://lermagazine.com/article/stress-fractures-lessons-from-military-research. Accessed March 11, 2021.

3. Almeida SA, Williams KM, Shaffer RA, Brodine SK. Epidemiological patterns of musculoskeletal injuries and physical training. Med Sci Sports Exerc 1999;31(8):1176–1182.

4. Caesar BC, McCollum GA, Elliot R, et al. Stress fractures of the tibia and medial malleolus. Foot Ankle Clin 2013;18(2):339–355.

5. McLaughlin R, Wittert G. The obesity epidemic: implications for recruitment and retention of defence force personnel. Obes Rev 2009 Nov;10(6):693–699.

6. Maxey H, Bishop-Josef S, Goodman B. Unhealthy and Unprepared: National Security Depends on Promoting Healthy Lifestyles from an Early Age. Washington, DC: Council For A Strong America, 2018.

7. Knapik JJ, Sharp MA, Montain SJ. Association between stress fracture incidence and predicted body fat in United States Army Basic Combat Training recruits. BMC Musculoskelet Disord 2018 May 22;19(1):161.

8. Fraser JJ, Schmied E, Rosenthal MD, Davenport TE. Physical therapy as a force multiplier: Population health perspectives to address short-term readiness and long-term health of military service members. Cardiopulm Phys Ther Journal 2020;31(1):22–28.

9. Waterman BR, Gun BK, Bader JO, et al. Epidemiology of lower extremity stress fractures in the United States military. Mil Med 2016;181(10):1308–1313.

10. MacGregor AJ, Han PP, Dougherty AL, Galarneau MR. Effect of dwell time on the mental health of US military personnel with multiple combat tours. Am J Public Health. 2012 Mar;102 Suppl 1(Suppl 1):S55–S59.

11. Hauret KG, Shippey DL, Knapik JJ. The physical training and rehabilitation program: duration of rehabilitation and final outcome of injuries in basic combat training. Mil Med 2001;166(9):820–826.

12. Reis JP, Trone DW, Macera CA, Rauh MJ. Factors associated with discharge during marine corps basic training. Mil Med 2007;172(9):936–941.

13. Roy TC, Faller TN, Richardson MD, Taylor KM. Characterization of limited duty neuromusculoskeletal injuries and return to duty times in the U.S. Army during 2017-2018. Mil Med 2021 Jan 9:usaa392. doi:10.1093/milmed/usaa392. Online ahead of print.

14. Cosman F, Ruffing J, Zion M, et al. Determinants of stress fracture risk in United States Military Academy cadets. Bone 2013 Aug;55(2):359–366.

15. Knapik J, Montain SJ, McGraw S, Grier T, Ely M, Jones BH. Stress fracture risk factors in basic combat training. Int J Sports Med. 2012 Nov;33(11):940–946.

16. U.S. Department of Defense, Office of the Under Secretary of Defense, Personnel and Readiness: Occupational conversion index: Enlisted/officer/civilian. DoD 1312.1-1 03/01/01. Available at https://apps.dtic.mil/dtic/tr/fulltext/u2/a327949.pdf. Accessed on November 17, 2020.

17. Burnes T. Contributions of women to U.S. combat operations. Carlisle Barracks, PA: U.S. Army War College, 2008. Available at: https://apps.dtic.mil/dtic/tr/fulltext/u2/a479020.pdf. Accessed November 17, 2020.

18. Sheppard C. Women in combat. Carlisle Barracks, PA: U.S. Army War College, 2007. Available at: https://apps.dtic.mil/dtic/tr/fulltext/u2/a467244.pdf. Accessed November 17, 2020.

19. Kamarck KN. Women in combat: Issues for congress. Washington, DC: Congressional Research Service, 2016. Available at: https://fas.org/sgp/crs/natsec/R42075.pdf. Accessed November 17, 2020.

20. Englert RM, Yablonsky AM. Scoping review and gap analysis of research related to the health of women in the U.S. military, 2000 to 2015. J Obstet Gynecol Neonatal Nurs 2019 Jan;48(1):5–15.

21. Armed Forces Health Surveillance Branch. Defense Medical Epidemiology Database (DMED) 5.0 Users Guide V. 1.0. March 2017. Available at: file:///C:/Users/Andrew/Downloads/DMEDUsersGuidever5.pdf. Accessed November 18, 2020.

22. Commission on Professional Hospital Activities. International Classification of Diseases, 9th Revision, Clinical Modification. Ann Arbor, MI: Edwards Brothers, 1977.

23. Glaviano NR, Boling MC, Fraser JJ. Anterior knee pain risk differs between male and female military tactical athletes. medRxiv 2020:2020.09.17.20196741. doi:10.1101/2020.09.17.20196741

24. Fraser JJ, MacGregor AJ, Ryans CP, Dreyer MA, Gibboney MD, Rhon DI. Sex and occupation are salient factors associated with lateral ankle sprain risk in military tactical athletes. J Sci Med Sport. 2021 Mar 4:S1440–2440(21)00052-9.

25. Dean AG, Sullivan KM, Soe MM. OpenEpi: Open Source Epidemiologic Statistics for Public Health, Version. April 2013. Available at: www.OpenEpi.com. Accessed December 18, 2020.

26. Belasco A. Troop levels in the Afghan and Iraq Wars, FY2001-FY2012: Cost and other potential issues. Washington, DC: Congressional Research Service, 2009. Available at: https://fas.org/sgp/crs/natsec/R40682.pdf. Accessed March 11, 2021.

27. Goldberg MS. Updated death and injury rates of U.S. military personnel during the conflicts in Iraq and Afghanistan. Working Paper 2014-08. Washington, DC: Congressional Budget Office, 2014. Available at: https://www.cbo.gov/sites/default/files/113th-congress-2013-2014/workingpaper/49837-Casualties_WorkingPaper-2014-08_1.pdf. Accessed March 11, 2021.

28. Carter LB. Iraq: summary of U.S. forces. Order Code RL31763. Washington, DC: Congressional Research Service, 2005. Available at: https://fas.org/sgp/crs/mideast/RL31763.pdf. Accessed March 11, 2021.

29. Bartlett JL, Phillips J, Galarneau MR. A descriptive study of the U.S. Marine Corps fitness tests (2000-2012). Mil Med 2015 May;180(5):513–517.

30. Hiebert R, Brennan T, Campello M, Lis A, Ziemke G, Faulkner D, et al. Incidence and mechanisms of musculoskeletal injuries in deployed Navy active duty service members aboard two U.S. Navy air craft carriers. Mil Med 2020 Sep 18;185(9-10):e1397–e1400.

31. Balcom TA, Moore JL. Epidemiology of musculoskeletal and soft tissue injuries aboard a U.S. Navy ship. Mil Med 2000 Dec;165(12):921–924.

32. Gregg MA 2nd, Jankosky CJ. Physical readiness and obesity among male U.S. Navy personnel with limited exercise availability while at sea. Mil Med 2012 Nov;177(11):1302–1307.

33. Rush T, LeardMann CA, Crum□Cianflone NF. Obesity and associated adverse health outcomes among US military members and veterans: Findings from the millennium cohort study. Obesity 2016;24(7):1582–1589.

34. Schuh-Renner A, Grier TL, Canham-Chervak M, Hauschild VD, Roy TC, Fletcher J, et al. Risk factors for injury associated with low, moderate, and high mileage road marching in a U.S. Army infantry brigade. J Sci Med Sport 2017 Nov;20 Suppl 4:S28–S33.

35. Birrell SA, Haslam RA. Subjective skeletal discomfort measured using a comfort questionnaire following a load carriage exercise. Mil Med 2009 Feb;174(2):177–182.

36. Orr RM, Pope R. Gender differences in load carriage injuries of Australian army soldiers. BMC Musculoskelet Disord 2016 Nov 25;17(1):488.

37. Omholt ML, Tveito TH, Ihlebæk C. Subjective health complaints, work-related stress and self-efficacy in Norwegian aircrew. Occup Med (Lond) 2017 Mar 1;67(2):135–142.

38. Nogueira HC, Diniz AC, Barbieri DF, Padula RS, Carregaro RL, de Oliveira AB. Musculoskeletal disorders and psychosocial risk factors among workers of the aircraft maintenance industry. Work 2012;41 Suppl 1:4801–4807.

39. Lovalekar M, Keenan KA, Beals K, Nindl BC, Pihoker AA, Coleman LC, et al. Incidence and pattern of musculoskeletal injuries among women and men during Marine Corps training in sex-integrated units. J Sci Med Sport 2020 Oct;23(10):932–936.

